# Projected Burden of Bloodstream Infections and the Impact of Molecular Rapid Diagnostic Testing in England and the United States (2025–2029)

**DOI:** 10.64898/2026.03.18.26348587

**Authors:** James K. Karichu, Mark Pennington, Kiera Lander, Tiffeny T. Smith, Adam Thornberg

## Abstract

**Introduction:** Data on bloodstream infections (BSI) indicate a growth in incidence over time. This analysis utilised national data from England and the best available United States (US) evidence to predict BSI incidence over the years 2025 to 2029. The analysis utilised evidence on the cost-effectiveness of molecular rapid diagnostic tests (mRDT) to estimate the cost and mortality associated with BSI, and the potential for increased use of mRDT to save lives.

**Methods:** Data on BSI incidence by age group and sex for England in 2017 and the US (Minnesota) for 2003 to 2005 were combined with demographic projections over the years 2025 to 2029 to estimate the number of BSIs. Published costs and mortality associated with BSI, according to the method of identification of the pathogen, were used to estimate the lives saved and the cost impact of widespread use of mRDT in England and the US.

**Results:** BSI cases in England and the US are predicted to total 1.02 million and 6.24 million over the years 2025 to 2029, associated costs are £14.6 million and $221 million, respectively. Expanding the use of mRDT would save 2,219 and 7,554 lives in England and the US, respectively, over a 5-year period and would reduce healthcare expenditure in both countries.

**Conclusion:** There is a compelling argument to increase the uptake of mRDT to improve patient outcomes. This analysis demonstrates that expanded mRDT adoption can significantly reduce BSI burden, saving over 9,700 lives and decreasing healthcare expenditure in both countries.

**Key messages:**

- There are an estimated 186,000 BSIs in England annually, with an associated mortality of around 21,000 deaths. In the US, there are over 1.12 million BSIs per year, with over 125,000 deaths.
- Over a five-year period in England, introduction of the Cobas® Eplex blood culture identification (BCID) panels would save 2,219 lives and would also reduce healthcare costs by over £50 million, while in the US, introduction of the Cobas Eplex BCID panels in hospitals not using mRDT would lead to over 7,500 lives saved and a cumulative saving of over $500 million over the period 2025 to 2029.
- Use of mRDT can save lives and reduce costs through more rapid identification of BSIs and faster optimisation of therapy.

## INTRODUCTION

Bloodstream infection (BSI) remain a major cause of morbidity and mortality among infectious diseases worldwide [1–3]. The substantial burden results in a significant financial toll on healthcare systems [1, 4–6]. However, there is surprisingly limited national surveillance data, particularly in the United States (US). In England, 2017 national data reported a BSI incidence of 248.7 cases per 100,000 population [7]. By contrast, robust national US incidence data are notably absent; the most widely cited population-based study, now nearly two decades old, reported an incidence of 154 cases per 100,000 in a single Minnesota county from 2003 to 2005 [8]. This lack of current, national-level data creates a significant evidence gap, obscuring the true, evolving burden of BSI and the potential impact of new diagnostic technologies.

Reducing BSI-related mortality and costs hinges on a single critical factor; timely and targeted antimicrobial therapy [5, 9]. Standard microbiological practice for pathogen identification can, however, introduce significant, clinically meaningful delays. Matrix-assisted laser desorption ionization-time of flight mass spectrometry (MALDI-ToF MS) has reduced time to pathogen identification, but typically requires overnight culturing and provides no information on antimicrobial resistance. Advanced MALDI-ToF MS techniques can avoid the need for culturing, but crucial susceptibility results typically require another full day [10]. This 24- to 48-hour diagnostic gap necessitates the use of empiric broad-spectrum antimicrobials, which may be ineffective, drive antimicrobial resistance, or increase healthcare costs [5, 11].

Molecular rapid diagnostic tests (mRDT) have changed the landscape to address this critical delay. By providing pathogen identification in hours rather than days, mRDT can optimise antimicrobial therapy, reduce costs, and subsequently improve clinical outcomes, including mortality rates [12]. We recently quantified the per-patient cost-effectiveness of various mRDT, demonstrating that Cobas® Eplex blood culture identification (BCID) panels were the most cost-effective mRDT [13].

While the previous analysis established per-patient value, it did not quantify the total, forward-looking national burden of BSI, particularly considering shifting population demographics. The present study addresses this critical gap. We utilise the best available data to project BSI incidence, mortality, and costs over the years 2025 to 2029 in the England and US. We then leverage parameters from our previous study [13] to estimate the total national-level impact that widespread mRDT adoption could have on saving lives and reducing healthcare expenditure.

## METHODS

### Overview

The study uses English national data on BSI incidence from 2017 as a function of age and sex, in conjunction with demographic projections to estimate BSI incidence in England over the years 2025 to 2029. The projections are combined with data from a recent analysis that compared the cost-effectiveness of adding a mRDT to MALDI-ToF MS and showed that the use of mRDT improves outcomes and reduce costs, primarily through reductions in length of stay [13]. This analysis evaluates the clinical benefits and healthcare costs of implementing mRDT alongside MALDI-ToF MS in hospitals currently utilising the latter alone. The mRDT is assumed to be the Cobas Eplex BCID panels as previous analysis has shown this system to be the most cost-effective mRDT [13]. The analysis also estimates BSI incidence in the US over the years 2025 to 2029 based on the best available US data and estimates the costs and benefits of achieving 100% uptake of mRDT.

### Current care in England

Current care in England for patients with a suspected BSI is assumed to align with recommendations from the National Institute for Health and Care Excellence (NICE) [14]. Evidence suggests that few hospitals in England use mRDT, and that the use of MALDI-ToF MS following overnight culturation is common but not universal [15]. We assumed that current protocols for pathogen diagnosis consists of MALDI-ToF MS following overnight culturation; this is likely to be conservative as some hospitals may still rely on traditional culturation only.

### Current care in the US

Recently published US guidelines recommend the use of mRDT, provided systems are in place to rapidly communicate test results to clinicians [16]. The use of mRDT is common in the US but many hospitals still rely on conventional culture and MALDI-ToF MS, with survey data from 2023 indicating that just 34% of laboratories in the US use an mRDT to diagnose BSI [17]. As uptake is likely to have increased since 2023, this analysis assumed that 50% of laboratories already use an mRDT. The remaining 50% were assumed to rely on MALDI-ToF MS alone. The market share of the most commonly used mRDTs were estimated from market research analysis undertaken by Roche, which synthesised data from public tenders, third-party market intelligence, and field-based intelligence. The market shares assumed are: Cobas Eplex BCID panels – 15%; BioFire BCID panels – 15%; BioFire BCID2 panels – 45%; Diasorin Verigene BCID panels – 15%; Accelerate PhenoTest BC kit – 10%.

### Uptake of mRDT

The analysis considers the hypothetical impact of the introduction of a mRDT, adjunct to MALDI-ToF MS, for identification of pathogens and common resistance mechanisms in cases of suspected BSI in hospitals currently not using an mRDT (and assumed to be using MALDI-ToF MS alone). For simplicity, hospitals adopting a mRDT are assumed to choose Cobas Eplex BCID panels. The Cobas Eplex BCID panels have the broadest pathogen coverage of commercially available mRDT and previous analysis demonstrated it to be the most cost-effective BCID mRDT in English and US settings [13]. Hence market share in England is assumed to change from 100% MALDI-ToF MS alone to 100% Cobas Eplex BCID panels in combination with MALDI-ToF MS. The assumption of 100% uptake is varied in scenario analysis. Analysis for the US assumed that Cobas Eplex BCID panels with MALDI-ToF MS replaced MALDI-ToF MS alone, with the market share for other mRDT remaining unchanged (Table 1 in the supplementary material).

**Table 1:**
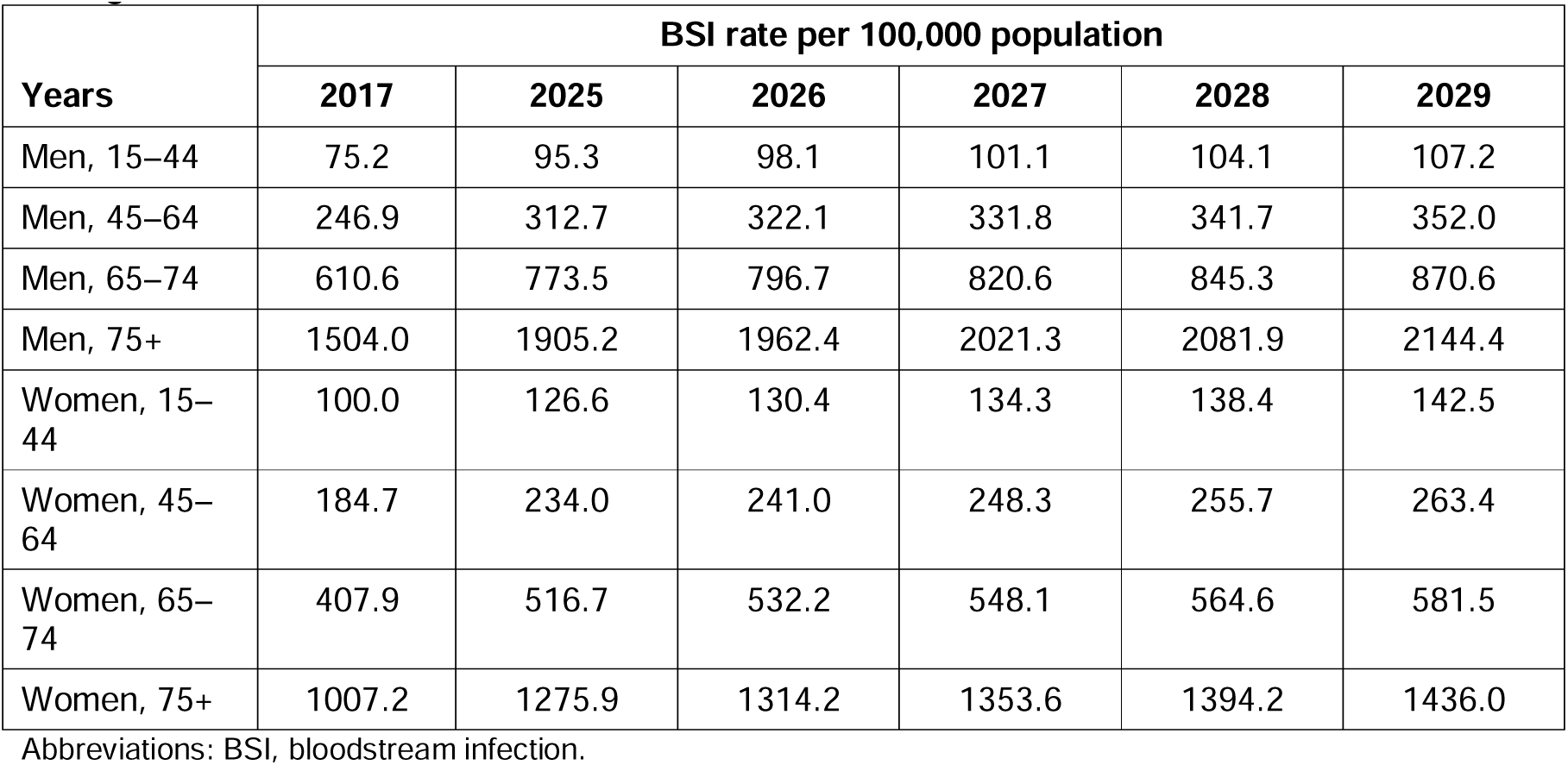
BSI incidence by age group and sex reported for 2017 and projected for 2025 to 2029 in England.

| Years | BSI rate per 100,000 population |  |  |  |  |  |
| --- | --- | --- | --- | --- | --- | --- |
|  | 2017 | 2025 | 2026 | 2027 | 2028 | 2029 |
| Men, 15–44 | 75.2 | 95.3 | 98.1 | 101.1 | 104.1 | 107.2 |
| Men, 45–64 | 246.9 | 312.7 | 322.1 | 331.8 | 341.7 | 352.0 |
| Men, 65–74 | 610.6 | 773.5 | 796.7 | 820.6 | 845.3 | 870.6 |
| Men, 75+ | 1504.0 | 1905.2 | 1962.4 | 2021.3 | 2081.9 | 2144.4 |
| Women, 15–44 | 100.0 | 126.6 | 130.4 | 134.3 | 138.4 | 142.5 |
| Women, 45–64 | 184.7 | 234.0 | 241.0 | 248.3 | 255.7 | 263.4 |
| Women, 65–74 | 407.9 | 516.7 | 532.2 | 548.1 | 564.6 | 581.5 |
| Women, 75+ | 1007.2 | 1275.9 | 1314.2 | 1353.6 | 1394.2 | 1436.0 |

Abbreviations: BSI, bloodstream infection.

The analysis assumes an antimicrobial stewardship programme (ASP) is in place, which provides 24-hour monitoring of test results and rapid communication of results to the treating clinician. ASPs are intended to provide guidance to clinicians on escalation or de-escalation of therapy with the aim of reducing adverse outcomes for patients and the development of antibiotic resistance [18]. Programme composition varies but typically includes a multidisciplinary team consisting of an infectious disease physician and a clinical pharmacist, along with infrastructure for the rapid communication of test results. There is evidence to indicate that early identification of pathogens has limited impact on patient care in the absence of an ASP [5].

### Incidence of BSIs - England

Estimates of the annual change in incidence of BSIs in the literature range from –3.3% to 5.2% per year. Evidence from England indicates a plateau in incidence of BSI rates between 2009 and 2014 with a sharp increase over the years 2014 to 2017 [7]. The combined rate of monomicrobial and polymicrobial BSI was 167.2 per 100,000 in 2009, rising to 248.7 per 100,000 in 2017, equivalent to an annual increase of 5.1%. National Infectious Disease Register data from Finland over the period 2004 to 2018 indicates a rise from 150 to 309 cases per 100,000 corresponding to an increase of 5.2% (95% CI: 4.8%, 5.5%) per year [2]. National surveillance data from Switzerland reported a BSI rate of 211 per 100,000 in 2011 rising to 240 per 100,000 in 2014, an annual increase of 4.4% [19]. Population data for Northern Denmark showed an increase in age- and sex-standardised BSI incidence from 114 per 10,000 population in 1992 to 166 per 100,000 in 2006, a 46% (95% CI: 36%, 59%) increase over the period, or 2.72% per year [20]. Sources consistently show a very large age gradient in the risk of BSI and hence some of the observed increases may have arisen from changing demographics and a growing proportion of elderly and very elderly patients over time. Consequently, we assumed that the rate of BSI is increasing at 3.0% annually in the base case. In scenario analysis we assume a rate of 5.0% per year.

The most recent data on BSI incidence in England were reported for 2017 and are broken down by age category and sex [7]. The rates for 2017 were combined with the estimated annual increase of 3.0% per year to estimate rates for the years 2025 to 2029 (Table 1).

Demographic data for the years 2025 to 2029 for England were taken from national population projections undertaken by the Office for National Statistics (ONS) [21]. The ONS produces population projections every 2 years based on mid-year population estimates, together with assumptions of future levels of fertility, mortality and migration, and represent the ‘best estimates’ of the future demographic composition of the UK nations. Population projections were combined with BSI rates by age group and sex to estimate the number of BSIs in England for the years 2025 to 2029.

### Incidence of BSIs - US

A review of the literature did not identify any national data on the incidence of BSI in the US. The relevant studies found were limited to specific populations, typically hospitalised patients. The best available evidence was a population-based study in Olmsted County, Minnesota over the period 2003 to 2005 [8]. The study reported a rate of 189 per 100,000 population. While this study is nearly two decades old, and limited to a single county, a population-based approach was favoured over more recent studies reporting hospitalised patient populations. Hospitalised patient cohorts likely overrepresent high-risk individuals and fail to capture the full community burden of BSI. Consequently, the BSI rate in the US in 2004 was assumed to be 189 per 100,000 population. Assuming a 3% increase in the rate of BSI (chosen as a conservative base case) over the period 2004 to 2017 generates an estimate of 277.6 per 100,000 in 2017. This value is a little higher than the estimate for England in 2017 of 248.7 per 100,000. The ratio of the estimated US rate in 2017 to the reported English rate for 2017 (1.116) was applied to BSI rates for England by age and sex category for the years 2025 to 2029 to estimate BSI rates for the US by age category over the same period (Table 2 in the supplementary material). Data on BSI rates were combined with data on the US population by age and sex group for the years 2025 to 2029 from the US Census Bureau [22], to estimate the number of BSIs in the US for the years 2025 to 2029. Extensive scenario analysis was conducted to assess the impact of assumptions and uncertainty in data sources including varying the annual growth rate of BSIs and varying the estimate of BSI derived from the Olmsted County study across the 95% confidence intervals reported.

**Table 2:**
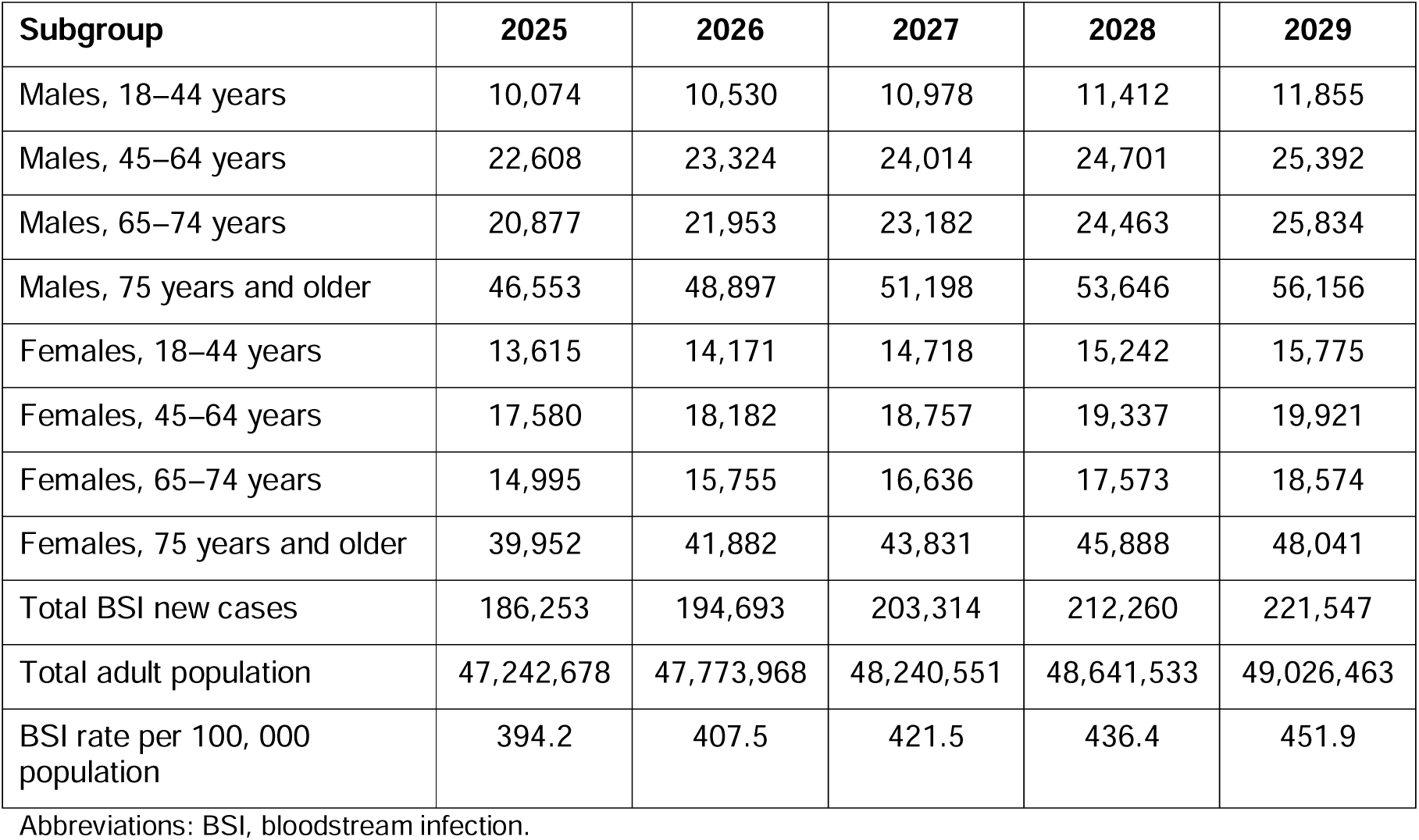
Predicted BSI incidence in England over the period 2025 to 2029.

### Impact of mRDT

The impact of adding mRDT to MALDI-ToF MS for detection of bloodstream pathogens and common resistance mechanisms was taken from a recent publication, which examined the cost-effectiveness of mRDT [13]. That analysis considered the impact on mortality and length of stay (LOS) of earlier identification of the pathogen responsible for BSI. The study considered reductions in LOS and mortality arising from reducing time to effective therapy in patients receiving ineffective broad-spectrum antimicrobial therapy, and reductions in adverse events (acute kidney injury [AKI] and *Clostridioides difficile* infection) from exposure to broad-spectrum antimicrobial therapy. The study reported the cost of a BSI and the number of patients surviving according to the mRDT used to supplement MALDI-ToF MS, in both an English and US setting. Data taken from the publication and applied in this analysis are reported in Table 3 of the supplementary material. Costs are reported for the price-year of 2024.

**Table 3:** Costs and mortality associated with BSI in England over the period 2025 to 2029 and the impact of mRDT.

|  | 2025 | 2026 | 2027 | 2028 | 2029 | Total over five years |
| --- | --- | --- | --- | --- | --- | --- |
| BSIs per year | 186,253 | 194,693 | 203,314 | 212,260 | 221,547 | 1,018,067 |
| Deaths per year under current care | 21,320 | 22,287 | 23,273 | 24,297 | 25,360 | 116,538 |
| Cost per year under current care | £2,669m | £2,790m | £2,913m | £3,042m | £3,175m | £14,589m |
| Lives saved per year with mRDT | 406 | 424 | 443 | 463 | 483 | 2,219 |
| Cost saved per year with mRDT | £9,312,645 | £9,734,661 | £10,165,711 | £10,613,012 | £11,077,334 | £50,903,363 |

Abbreviations: BSI, bloodstream infection; m, million; mRDT molecular rapid diagnostic test.

### Scenario analysis

The following scenario analyses were run to explore uncertainty in key parameters:

- Annual increase in the underlying risk of BSI of 1.0% and 5.0%.
- BSI rate for the US in 2004 of 174 and 204 based on the upper and lower confidence intervals of the estimate from the Minnesota study [8].
- Linear increase in uptake of mRDT from 60% in 2025 to 100% in 2029 in the US, and from 20% in 2025 to 100% in 2029 in England.
- The proportion of hospitals in the US considered to be using mRDT in addition to MALDI-ToF MS varied between 40% and 60%.
- Mortality rates 10% higher or lower than the base case.
- A cost saving of £55 and £45 associated with the addition of the Cobas Eplex BCID panels to MALDI-ToF MS in an English setting, representing values 10% higher or lower than the base case, respectively.
- A cost saving of $90.20 and $73.80 associated with the addition of the Cobas Eplex BCID panels to MALDI-ToF MS in a US setting representing values 10% higher or lower than the base case, respectively.

## RESULTS

The projected BSI incidence for England over the five years from 2025 to 2029 is shown in Table 2. The number of BSIs was predicted to rise from 186,253 in 2025 to 221,547 in 2029, equating to a rise from 394.2 to 451.9 per 100,000 population. Figure 1 shows the BSI rate per 100,000 population under the base case assumption of 3% growth per year and in scenarios assuming 1% and 5% growth per year, for both England and the US. Predictions for the US are around 12% higher than for England, reflecting the ratio of the predicted number of BSIs in the US in 2017 to the observed number of BSIs in England in 2017. Predicted numbers of BSIs in 2025 are around 35% higher under the scenario of 5% growth compared with the scenario of 1% growth; by 2029 predictions diverge by approximately 60% across the two scenarios. This highlights the critical impact of both demographic shifts and underlying incidence trends on the future BSI burden.

**Figure 1:**
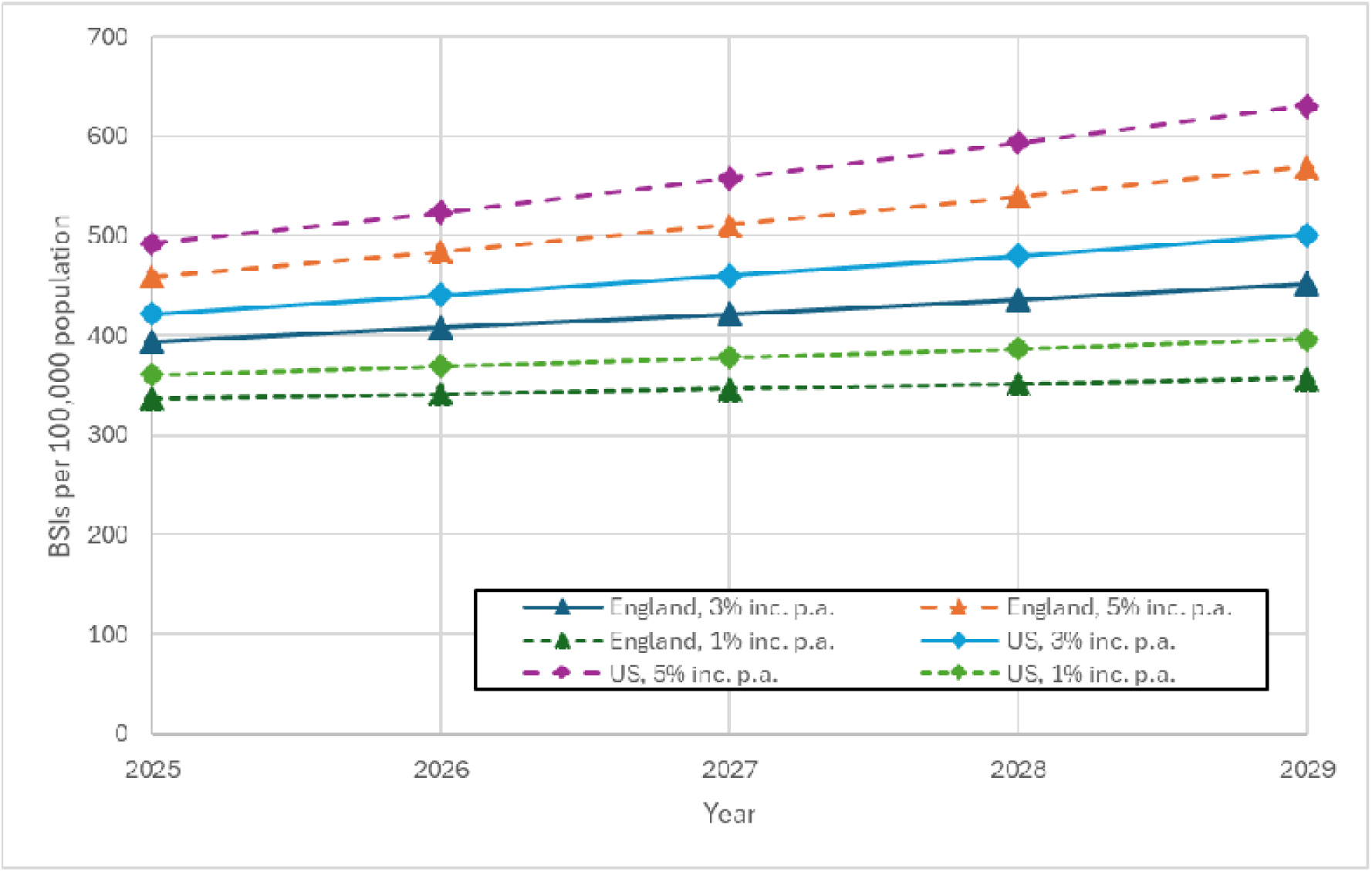
BSI rates for the period 2025 to 2029 in the base case and scenarios for England and US. Abbreviations: BSI, bloodstream infection; p.a., per annum; US, United States.

The costs and mortality associated with BSIs in England over the years 2025 to 2029 are reported in Table 3. There are 21,320 deaths predicted as a result of BSI in 2025 rising to 25,360 in 2029. Over the five-year period, 116,538 deaths are predicted as a result of BSIs, and the cost is £14.6 million. Introduction of the Cobas Eplex BCID panels would save 406 lives in 2025, rising to 483 in 2029. Over the five-year period, introduction of the Cobas Eplex BCID panels would save 2,219 lives and would also reduce healthcare costs by £51 million. Implementing mRDTs reduces overall five-year BSI mortality by 1.9%. These gains, alongside cost savings that offset mRDT acquisition, are driven by shorter times to effective therapy and decreased exposure to broad-spectrum antimicrobials, which mitigates risks of AKI, *C. difficile* infection, and BSI-associated length of stay (LOS). Figure 2 shows the projected number of BSIs over the years 2025 to 2029 and the number of lives saved each year through the use of mRDT in the base case and in the scenario in which BSI incidence rises at 5% per year.

**Figure 2:**
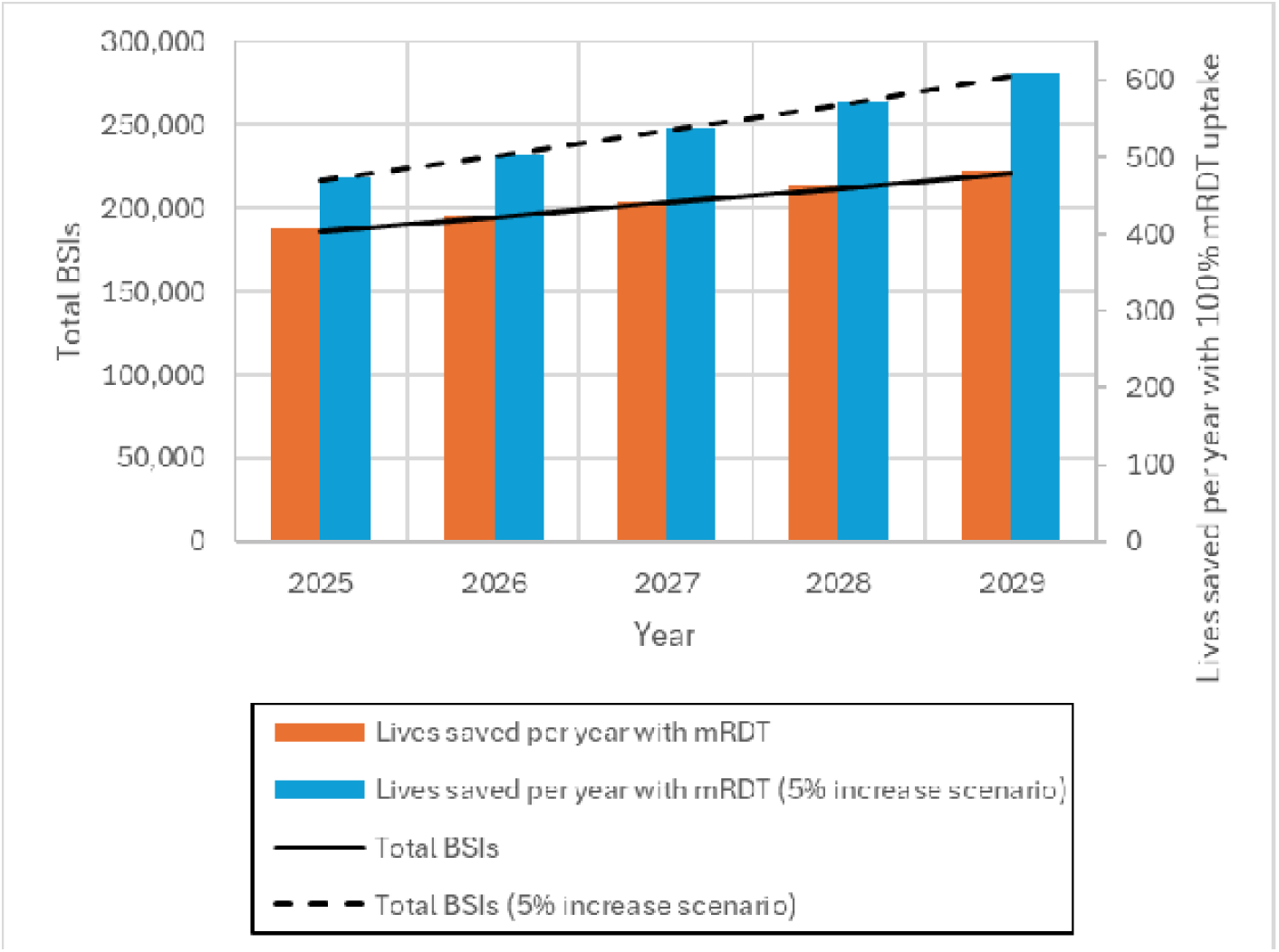
Number of BSIs and the number of lives saved by adoption of mRDTs over the years 2025 to 2029 in the base case and scenario assuming a higher rate of annual increase in BSI. Abbreviations: BSI, bloodstream infection; mRDT, molecular rapid diagnostic test.

### BSI incidence, costs and lives saved by mRDT uptake in the US

The projected BSI incidence by age and sex for the US over the years 2025 to 2029 is shown in Table 4 in the supplementary material. There is a sharp increase in the number of BSIs amongst those aged over 75 years, driven by demographic growth. The overall number of BSIs is predicted to grow by 22% over the five years from 1,126,813 to 1,374,519 due to a combination of rising BSI incidence and demographic changes. The overall BSI rate is predicted to rise from 421.9 to 500.8 per 100,000 population over the five years.

**Table 4:**
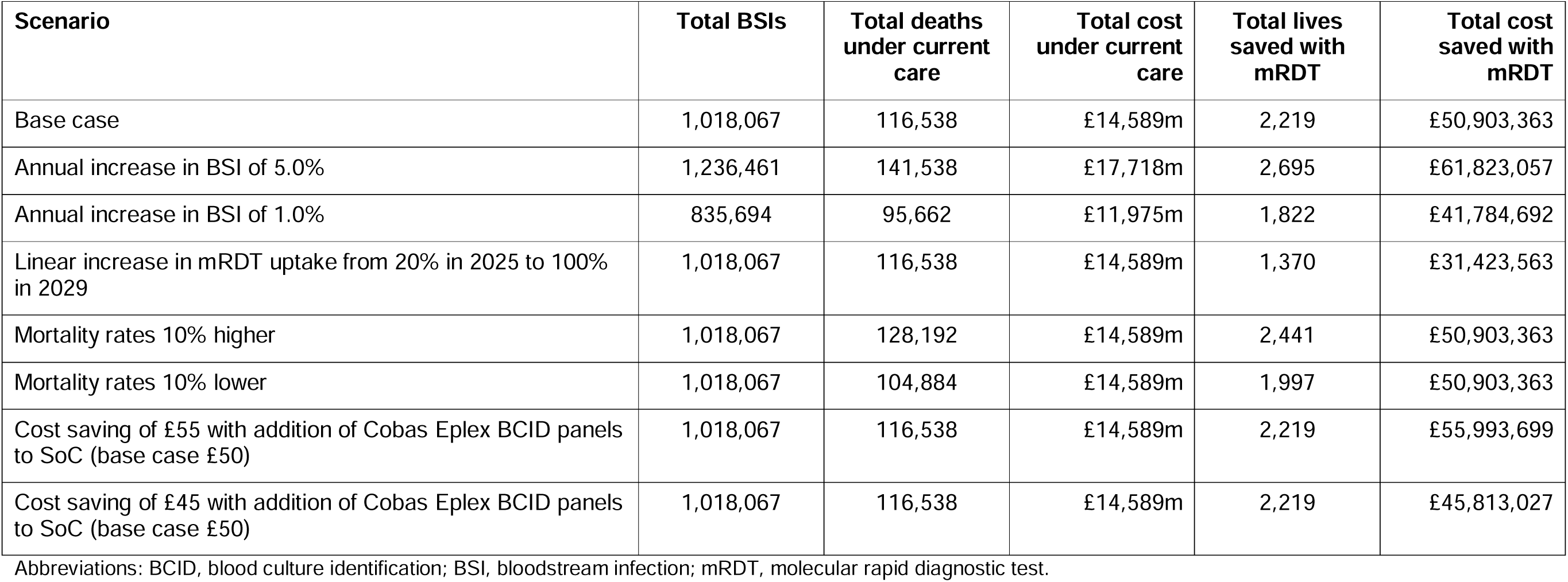
Key results from the scenario analyses.

| **Scenario** | **Total BSIs** | **Total deaths under current care** | **Total cost under current care** | **Total lives saved with mRDT** | **Total cost saved with mRDT** |
| --- | --- | --- | --- | --- | --- |
| Base case | 1,018,067 | 116,538 | £14,589m | 2,219 | £50,903,363 |
| Annual increase in BSI of 5.0% | 1,236,461 | 141,538 | £17,718m | 2,695 | £61,823,057 |
| Annual increase in BSI of 1.0% | 835,694 | 95,662 | £11,975m | 1,822 | £41,784,692 |
| Linear increase in mRDT uptake from 20% in 2025 to 100% in 2029 | 1,018,067 | 116,538 | £14,589m | 1,370 | £31,423,563 |
| Mortality rates 10% higher | 1,018,067 | 128,192 | £14,589m | 2,441 | £50,903,363 |
| Mortality rates 10% lower | 1,018,067 | 104,884 | £14,589m | 1,997 | £50,903,363 |
| Cost saving of £55 with addition of Cobas Eplex BCID panels to SoC (base case £50) | 1,018,067 | 116,538 | £14,589m | 2,219 | £55,993,699 |
| Cost saving of £45 with addition of Cobas Eplex BCID panels to SoC (base case £50) | 1,018,067 | 116,538 | £14,589m | 2,219 | £45,813,027 |

Abbreviations: BCID, blood culture identification; BSI, bloodstream infection; mRDT, molecular rapid diagnostic test.

Predicted mortality and costs over the period 2025 to 2029 in the US are reported in Table 5 in the supplementary material. Deaths rise from 125,856 in 2025 to 153,522 in 2029, amounting to 697,289 over five years. Costs over the same period rise from $39.9 billion to $48.7 billion (2024 USD). Introduction of the Cobas Eplex BCID panels in hospitals relying only on MALDI-ToF MS would save 1,363 lives in 2025, increasing to 1,663 in 2029. Over 7,500 lives would be saved over the five-year period. The cost impact of supplementing MALDI-ToF MS with the Cobas Eplex BCID panels in hospitals not using mRDT would be a saving of $92.4 million in 2025 and a cumulative saving of $512 million over the period 2025 to 2029.

### Results of scenario analyses

Table 4 reports results of the scenario analyses. The scenarios applying a 1% or a 5% increase in BSI per annum generated a notable impact on overall costs, deaths attributable to BSI and the benefits of mRDT; outcomes were roughly 20% higher in the scenario assuming a 5% increase and correspondingly 20% lower in the scenario assuming a 1% increase. The scenario assuming a linear increase in uptake of mRDT from 20% in 2025 to 100% in 2029 generated the lowest estimate of costs saved and lives saved across all English scenarios. However, at 1,370, the lives saved in this scenario remained substantial. Scenarios varying cost and mortality inputs by ±10% generated results in which cost savings or lives saved by the uptake of mRDT also varied by ±10%. Overall, the scenario analyses demonstrate that while the magnitude of the benefit fluctuates with different inputs, the conclusion—that widespread mRDT adoption saves thousands of lives and is cost-saving— remains robust across all tested variations.

The results of the scenario analyses in a US setting are provided in Table 6 of the supplementary material. The sensitivity of the results in the US setting to changes in input parameters was in line with observations for the England analysis. The scenarios in which the annual rate of increase in BSI was assumed to increase at 1% and at 5% a year generated results which were roughly 20% lower or higher than the base case, respectively. The lowest number of lives saved and costs saved were observed in the scenario in which uptake of mRDT increases linearly each year to 100% in 2029. Varying cost savings or reduction in mortality associated with mRDT by 10% had a similar proportionate impact on the related cost or mortality output. Increasing or decreasing the proportion of hospitals relying on MALDI-ToF MS alone by 20% generated a proportionate increase or decrease in lives saved and costs saved with 100% uptake of mRDT. Across all scenarios, lives saved and cost savings associated with the uptake of mRDT were substantial with a minimum of 4,682 lives saved and $317 million costs saved.

## DISCUSSION

### Key findings

This analysis provides, to our knowledge, the first forward looking projection of the significant and growing BSI burden in England and the US, estimating over 1.0 million and over 6.2 million cumulative cases in these geographies, respectively, by 2029. Our findings quantify the substantial human toll, with over 115,000 projected deaths in England and nearly 700,000 projected deaths in the US, and demonstrate the potential for mRDT to save thousands of lives. This burden is driven not only by population growth but by a projected increase in incidence among the most vulnerable, elderly populations. After assuming an increase in incidence of BSI of 3.0% annually since 2017, incidence of BSIs in England is predicted to rise from 394 to 452 per 100,000 population. For the US, our analysis predicts a rise from 422 to 501 per 100,000 population over the five-year period. The increased rate over time is partly a consequence of demographic changes, which are more marked in the predictions for the US.

Implementing existing mRDT technology could prevent approximately 2,200 deaths in England and 7,500 deaths in the US. Recent analysis has shown that such technology is not only cost-effective, but cost saving compared with conventional culture and MALDI-ToF MS alone, with reported savings of $164 and £50 per patient receiving an mRDT with the Cobas Eplex BCID panels compared with MALDI-ToF MS alone [13]. This analysis shows that widespread use of mRDT could save the National Health Service (NHS) £50 million over five years and could reduce US health care costs by $512 million over the same period. This analysis moves beyond a static, per-patient cost-effectiveness calculation (14) by quantifying the total national-level opportunity. We provide a framework for policymakers and hospital administrators to visualise the human and economic cost of diagnostic inertia over the next five years.

Evidence from a systematic review indicates similar incidence of BSI and associated mortality rates in England, the US, and other developed countries [23]. Hence it is likely that the relative magnitude of mortality gains following uptake of mRDT (1.9% in England) are generalisable to other developed countries in which current uptake of mRDT is low. The impact of mRDT on healthcare costs is dependent on the trade-off between test costs and cost savings from shorter LOS, and may be less generalisable.

### Interpretation of the results

This analysis illustrates the potential for mRDT to improve patient outcomes without increasing healthcare budgets. The high-profile failure to diagnose sepsis, which caused the death of Martha Mills, led to changes in care in England and the right of healthcare staff or family members to request a second opinion on a sepsis diagnosis if they believe the condition of the patient is declining [24]. While such changes have the potential to save lives, implementation is dependent on staff availability and the willingness of staff or the patient’s relatives to advocate for the patient. The ruling also highlights the challenges of diagnosis based on clinical presentation in the absence of definitive data on infections. Rapid identification of bloodstream pathogens and common resistant mechanisms can sidestep these challenges, allowing optimisation of therapy before the signs of patient decline become apparent. Implementation of mRDT in England has the potential to save hundreds of lives a year without leading to increased healthcare costs or demands on frontline staff time.

Previous NICE assessment of the procalcitonin tests for sepsis in 2015 failed to lead to a positive recommendation, despite evidence that such tests reduce both mortality and costs [25]. The guidance noted the potential for the technology to save lives but considered the available evidence insufficient to make a positive recommendation. Generating clinical trial evidence of improved mortality to justify a very modest test cost is extremely challenging. Reductions in mortality of the order of 1 in 500 patients, which are more than sufficient to justify the cost of the test, would require a very large trial. Consequently, definitive trial data may never be available. Decision models allow synthesis of data on intermediate outcomes, such as time to effective therapy and the relationship with mortality, to estimate final outcomes in circumstances where generating evidence is impractical [26]. Waiting for definitive data on the impact of mRDT on mortality may postpone a decision indefinitely. Decisions on implementation should be made following synthesis of the best available data when definitive data are unlikely to become available.

### Strengths and limitations

This analysis updates estimates of BSI incidence for England and US previously reviewed in 2013 [23]. The analysis used national laboratory surveillance data for England, which provided granularity on the incidence of BSI by age and sex. We examined different plausible scenarios for the growth in BSI rate over time, and applied the adjusted rates to demographic data for England for the period 2025 to 2029 to generate an overall BSI rate reflecting changes in population demographics.

Data for the US on BSI incidence relied on a study from Olmsted County in Minnesota [8]. Estimates also assumed that the relative risk of BSI as a function of age and sex is the same as for England. Age is a strong predictor of BSI rate. However, public coverage of healthcare costs as a function of age (MediCare) in the US, but not England, might drive differences in the age gradient. Whilst our analysis quantified the impact of using the upper and lower confidence intervals of the BSI rates estimated for Olmsted, Minnesota, this may be insufficient to reflect the extent to which the BSI rate in Olmsted differs from the average across the US. The median age of the population of Olmsted was 1.5 years less than the overall US population in 2023 [27]. Lower socio-economic status has also been liked to increased BSI incidence [28]. Median household income in Olmsted was 10% higher than the US average in 2023 [27]. There is also evidence of increased incidence of BSI with higher outdoor temperatures [29]. Hence it seems likely that the BSI rate in Olmsted would have been lower than the average for the US in 2004. Nonetheless, inflating the underlying rate from 2004 to 2017 by 3.0% a year generated a value broadly aligned with English data for 2017.

The data on effectiveness of mRDT used in this analysis includes an assumption of round-the-clock availability of pathology services and the infrastructure to convey results to treating clinicians with guidance on optimisation of antimicrobial therapy, where required. These components of an ASP are recognised as a central aspect in efforts to reduce antimicrobial resistance, as well as optimising outcomes for patients. Our analysis demonstrates the gains that mRDT can provide when complementing an effective ASP.

This analysis builds on a recent cost-effectiveness analysis of mRDTs which sought to synthesise the best available evidence on the impact of mRDT on costs and mortality [13]. A key feature of that analysis was the use of data on the effectiveness of empiric therapy across different pathogens, and the subsequent realistic appraisal of the potential of rapid testing to improve mortality for patients receiving ineffective empiric therapy. While that analysis provides a robust estimate of the lives saved through the application of mRDT to reduce time to effective therapy for BSIs, the overall mortality rate estimated from that analysis is likely to be an underestimate as the analysis included contamination results for which a mortality of zero was assumed. There are further limitations in using this data to estimate the overall mortality from BSIs, as the study did not adjust mortality estimates for age. Consequently, the estimates of the number of deaths in this analysis do not account for ageing populations. For these reasons, the mortality estimates in this study should be considered a lower estimate of the true number of deaths. Consequently, the numbers of lives lost to BSI, and the potential for mRDT to save lives, presented here are likely to be conservative.

Data on the uptake of mRDT in England and the US is limited and our estimates are subject to uncertainty. We conservatively assumed that 50% of microbiology laboratories in the US are already using mRDT. This would represent a sizeable increase on the datum for 2023. A lower current uptake would translate into larger mortality gains and cost savings than we have calculated. Our estimates of the mortality and cost impact of introducing mRDT are dependent on a previously published cost-effectiveness analysis. Further analysis of the cost-effectiveness of mRDT would help to refine estimates of the costs and benefits. Our base case analysis demonstrates the scope for mRDT to save lives with full implementation in 2025. A staged increase in the uptake of mRDT may be more feasible. Costs and lives saved fall under scenarios assuming a linear increase in uptake to 100% in 2029 but remain substantial.

## Conclusion

Bloodstream infections pose a large and growing public health challenge. Our analysis projects 1.0 million cases and 116,000 deaths in England, and 6.2 million cases and nearly 700,000 deaths in the US, over the period 2025 to 2029. This increasing burden is partly driven by demographic changes. The impact of rising BSI incidence can be partly offset by the widespread use of mRDT. This study demonstrates the potential for mRDT to save over 2,200 lives in England and more than 7,500 lives in the US, while generating cumulative healthcare savings of $512 million and £50 million, respectively, from 2025 to 2029. The use of mRDTs has been shown to be cost-effective, and even cost-saving, compared with traditional microbiology methods alone. Beyond direct clinical and economic gains, widespread mRDT adoption aligns with critical patient safety priorities and strengthens ASPs. Reappraisal of the role of mRDT is merited on the strength of the available evidence.

## DECLARATIONS

## Data Availability

The analysis used secondary data sources which are publicly available. No primary individual patient data was collected or used.
The model is considered commercially sensitive and we are unable to provide access to it.

## Acknowledgements

Roche would like to acknowledge Dr Dom Partridge at Source Health Economics for their medical writing and editorial support.

## Funding

COBAS and EPLEX are trademarks of Roche. All other product names and trademarks are the property of their respective owners.

Financial support for this research analysis and medical writing support was provided by GenMark Diagnostics (A member of the Roche Group) to Source Health Economics— content experts (MP).

## Conflicts of interest

JKK and AT are employees of Roche Molecular Systems (RMS). JKK and AT hold Roche stock options.

TS is an employee of Roche Diagnostics Corporation (RDC).

## Availability of data and material

The analysis used secondary data sources which are publicly available. No primary individual patient data was collected or used.

## Ethics approval

The analysis used secondary data sources. No individual patient data was collected or used. Consequently, ethical approval for the study was not required.

## Consent to participate

No individual patient data was collected or used, and hence consent was not required.

## Code availability

The model is considered commercially sensitive and we are unable to provide access to it.

## Author contributions

JKK and AT conceived the study. MP, JKK, AT and KL designed the study. MP, JKK, AT, and TS compiled the data. MP, JKK, AT, TS, and KL reviewed the results. MP drafted the manuscript. MP, JKK, AT, TS, and KL reviewed the manuscript.

