## Supplementary material for "Projected Burden of Bloodstream Infections and the Impact of Molecular Rapid Diagnostic Testing in England and the United States (2025–2029)"

Data inputs for the US analysis and data derived from a previous published analysis

Market shares for mRDT in the US

Table 1: Proportion of hospitals using mRDT currently and under hypothetical scenario in the US

|  | **MALDI-ToF MS alone** | **Cobas**® **Eplex BCID panels** | **BioFire BCID panel** | **BioFire BCID2 panel** | **Diasorin Verigene BCID panels** | **Accelerate PhenoTest BC kit** |
| --- | --- | --- | --- | --- | --- | --- |
| Status quo | 50% | 7.5% | 7.5% | 22.5% | 7.5% | 5% |
| Widespread adoption of mRDT | 0% | 57.5% | 7.5% | 22.5% | 7.5% | 5% |

Percentages represent the estimated percentage of microbiology laboratories or hospitals utilizing the specified diagnostic technology, based on 2023 survey data and market intelligence. For instance, it is assumed that 50% of the US market currently uses mRDTs (distributed across brands like BioFire, Cobas Eplex BCID panels, etc.), while the other 50% relies on MALDI-ToF MS alone. Widespread adoption assumes 50% of hospitals currently utilizing MALDI-ToF MS alone will adopt Cobas Eplex BCID panels.
Abbreviations: BCID, blood culture identification panel; MALDI- ToF MS, matrix-assisted laser desorption/ionisation - time of flight; mRDT molecular rapid diagnostic test; US, United States of America.

Projected BSI incidence by age group and sex for 2025 to 2029 in the US

Table 2: BSI incidence by age group and sex for 2025 to 2029 in the US

|  | **BSI incidence per 100,000 population** | | | | |
| --- | --- | --- | --- | --- | --- |
|  | **2025** | **2026** | **2027** | **2028** | **2029** |
| Men, 15–44 | 106.3 | 109.5 | 112.8 | 116.2 | 119.7 |
| Men, 45–64 | 349.0 | 359.5 | 370.2 | 381.3 | 392.8 |
| Men, 65–74 | 863.3 | 889.2 | 915.8 | 943.3 | 971.6 |
| Men, 75+ | 2,126.3 | 2,190.1 | 2,255.8 | 2,323.4 | 2,393.1 |
| Women, 15–44 | 141.3 | 145.5 | 149.9 | 154.4 | 159.0 |
| Women, 45–64 | 261.2 | 269.0 | 277.1 | 285.4 | 294.0 |
| Women, 65–74 | 576.6 | 593.9 | 611.7 | 630.1 | 649.0 |
| Women, 75+ | 1,423.9 | 1,466.6 | 1,510.6 | 1,555.9 | 1,602.6 |

Abbreviations: BSI, bloodstream infection.

Data on costs and mortality as a function of mRDT derived from published data

Table 3: Data on the cost and mortality impact of mRDT derived from Karichu et al. 2026

|  | **MALDI-ToF MS** | **Cobas Eplex BCID panels** | **BioFire BCID panel** | **BioFire BCID2 panel** | **Diasorin Verigene BCID panels** | **Accelerate PhenoTest BC kit** |
| --- | --- | --- | --- | --- | --- | --- |
| UK, cost of BSI episode | £14,330 | £14,280 | ̶† | ̶† | ̶† | ̶† |
| UK, mortality associated with BSI (per 10,000 patients) | 1,144.7 | 1,122.9 | ̶† | ̶† | ̶† | ̶† |
| US, cost of BSI episode | $35,515 | $35,351 | $35,372 | $35,364 | $35,365 | $35,456 |
| US, mortality associated with BSI (per 10,000 patients) | 1127.8 | 1103.6 | 1105.9 | 1104.7 | 1111.1 | 1108.3 |

†Only data for MALDI- ToF MS alone, and the Cobas Eplex BCID panels as an adjunct to MALDI-ToF MS were published for the UK. The UK analysis considers a transition from 100% MALDI-ToF MS alone to 100% Cobas Eplex BCID panels as an adjunct to MALDI- ToF MS as this was the most cost-effective mRDT (see Table 2).
Abbreviations: BCID, blood culture identification panel; BSI, bloodstream infection; MALDI- ToF MS, matrix-assisted laser desorption/ionisation - time of flight; mRDT molecular rapid diagnostic test; UK, United Kingdom; US, United States of America. Results in a US setting

Predicted BSI numbers in the US over the period 2025 to 2029

Table 4: Predicted BSI incidence in the US over the period 2025 to 2029

| **Subgroup** | **2025** | **2026** | **2027** | **2028** | **2029** |
| --- | --- | --- | --- | --- | --- |
| Males, 18–44 years | 66,141 | 68,416 | 70,654 | 72,921 | 75,262 |
| Males, 45–64 years | 141,211 | 145,079 | 149,218 | 153,589 | 158,028 |
| Males, 65–74 years | 146,580 | 153,975 | 161,107 | 167,998 | 174,626 |
| Males, 75 years and older | 246,977 | 265,569 | 285,416 | 306,167 | 328,114 |
| Females, 18–44 years | 84,881 | 87,816 | 90,703 | 93,625 | 96,630 |
| Females, 45–64 years | 107,151 | 110,017 | 113,088 | 116,321 | 119,617 |
| Females, 65–74 years | 109,335 | 114,569 | 119,635 | 124,518 | 129,163 |
| Females, 75 years and older | 224,539 | 240,120 | 256,764 | 274,361 | 293,079 |
| Total BSI new cases | 1,126,813 | 1,185,562 | 1,246,585 | 1,309,501 | 1,374,519 |
| Total adult population | 267,081,433 | 269,162,566 | 271,046,426 | 272,803,848 | 274,438,223 |
| BSI rate per 100,000 population | 421.9 | 440.5 | 459.9 | 480.0 | 500.8 |

Abbreviations: BSI, bloodstream infection.

Predicted mortality and costs attributable to BSI in the US over the period 2025 to 2029

Table 5: Costs and mortality associated with BSI in the US over the period 2025 to 2029 and the impact of mRDT

|  | **2025** | **2026** | **2027** | **2028** | **2029** | **Total** |
| --- | --- | --- | --- | --- | --- | --- |
| BSIs per year | 1,126,813 | 1,185,562 | 1,246,585 | 1,309,501 | 1,374,519 | 6,242,980 |
| Deaths per year under current care | 125,856 | 132,418 | 139,233 | 146,260 | 153,522 | 697,289 |
| Cost per year under current care | $39,939m | $42,021m | $44,184m | $46,414m | $48,718m | $221,275m |
| Lives saved per year with mRDT | 1,363 | 1,435 | 1,508 | 1,584 | 1,663 | 7,554 |
| Cost saved per year with mRDT | $92,398,683 | $97,216,118 | $102,219,947 | $107,379,042 | $112,710,576 | 511,924,365 |

Abbreviations: BSI, bloodstream infection; mRDT molecular rapid diagnostic test.

Scenario analysis in a US setting

Table 6: Results of scenario analyses in a US setting

| **Scenario** | **Total BSIs** | **Total deaths under current care** | **Total cost under current care** | **Total lives saved with mRDT** | **Total cost saved with mRDT** |
| --- | --- | --- | --- | --- | --- |
| Base case | 6,242,980 | 697,289 | $221,275m | 7554 | $511,924,365 |
| Annual increase in BSI of 5.0% | 7,584,049 | 847,076 | $268,808m | 9177 | $621,892,048 |
| Annual increase in BSI of 1.0% | 5,123,362 | 572,237 | $181,591m | 6199 | $420,115,723 |
| BSI rate of 204 in 2004 (upper CI from Uslan 2007) | 6,738,455 | 752,630 | $238,836m | 8154 | $552,553,283 |
| BSI rate of 174 in 2004 (lower CI from Uslan 2007) | 5,747,505 | 641,949 | $203,713m | 6954 | $471,295,447 |
| Linear increase in mRDT uptake from 60% in 2025 to 100% in 2029 | 6,242,980 | 697,289 | $221,275m | 4682 | $317,311,961 |
| Mortality rates 10% higher | 6,242,980 | 767,018 | $221,275m | 8309 | $511,924,365 |
| Mortality rates 10% lower | 6,242,980 | 627,560 | $221,275m | 6799 | $511,924,365 |
| Current proportion of hospitals using mRDT is 40% | 6,242,980 | 698,648 | $221,364m | 9065 | $614,309,238 |
| Current proportion of hospitals using mRDT is 60% | 6,242,980 | 695,931 | $221,186m | 6043 | $409,539,492 |
| Cost saving of $90.20 with addition of Cobas Eplex BCID panels to SoC (base case $82) | 6,242,980 | 697,289 | $221,275m | 7554 | $563,116,801 |
| Cost saving of $73.80 with addition of Cobas Eplex BCID panels to SoC (base case $82) | 6,242,980 | 697,289 | $221,275m | 7554 | $460,731,928 |

Abbreviations: BCID, blood culture identification; BSI, bloodstream infection; mRDT, molecular rapid diagnostic test.
